# Individual Symptom Severity in Adult Autism Spectrum Disorder Predicts Reduction in Right Dorsal Stream Neural Activity During Live Eye-to-Eye Contact

**DOI:** 10.1101/2021.10.07.21264650

**Authors:** Joy Hirsch, Xian Zhang, J. Adam Noah, Adam Naples, Julie M. Wolf, James C. McPartland

## Abstract

**Background:** Social symptomatology quantified by clinical interview (Autism Diagnostic Observation Schedule, ADOS) and self-report (Social Responsiveness Scale, SRS) indicate symptom severity in autism spectrum disorder (ASD). Reluctance to engage in interpersonal eye contact is a frequently observed behavioral hallmark, though neural bases for these difficulties and relation to symptomatology are not understood. We test the hypothesis that eye contact in ASD activates atypical neural mechanisms that are related to individual differences in symptomatology.

**Methods:** Neural activity represented by hemodynamic signals was acquired by functional near-infrared spectroscopy (fNIRS) during real person-to-person eye contact (confirmed by eye-tracking) for 17 adult ASD (3 female, 14 male) and 19 typically-developed (TD) participants (8 female, 11 male). Assessment of social function was based on ADOS scores for ASD participants and SRS scores for the combined group of ASD and TD participants.

**Results:** Individual ADOS scores were negatively correlated (r = -0.69) with individual fNIRS beta-values (representing strength of hemodynamic signals) within clusters in the right dorsal stream regions: somatosensory cortices, angular gyrus, and supramarginal gyrus. Hemodynamic responses in the right dorsolateral prefrontal cortex (DLPFC) were also negatively correlated (r = -0.77) with ADOS scores. Similarly, SRS scores for the combined ASD and TD groups were also negatively correlated (r = -0.58) with somatosensory cortices and the supramarginal gyrus.

**Conclusions:** These findings are consistent with the hypothesis that neural mechanisms in the dorsal stream and DLPFC are related to social symptomatology and implicate high-level interactive face and eye-processing systems as potential neurobiological markers of ASD.

## Introduction

Humans are highly sociable and typically endowed with the ability to rapidly perceive complex dynamic information conveyed by facial expressions. Faces are a primary source of social information, and the cues extracted from faces during interpersonal interactions guide dynamic and reciprocal perceptions and behaviors. Interpreting and reacting to faces is subserved by widely distributed neural networks consisting of functionally connected loci within early and high-level visual systems. Topographical maps associated with regional specializations for coding faces are well-established. For example, a region in the ventral-occipital cortex referred to as the “fusiform face area” is highly selective for and sensitive to faces (Haxby et al., 2000; Kanwisher et al., 1997). Regions within the superior temporal sulcus are involved in detecting dynamic facial movements (Allison et al., 2000; Puce et al., 1998), and elaborate face processing codes have been identified by primate electrophysiology in middle and superior temporal gyri (Chang & Tsao, 2017).

Due to the pre-eminence of face stimuli for social interactions and the innate and widespread biological specializations for face processing in the human brain, face-related tasks provide a very useful probe for assessing neural organization in developmental conditions, such as autism spectrum disorder (ASD). Difficulties with face-processing are well-documented in ASD, though associated differences in neural function have not yet been fully explained. For example, a relationship between social symptomatology and face processing is often observed, but mechanistic clarity remains an investigational goal (Golarai et al., 2006). Eye contact is a special case of interactive face processing particularly relevant in ASD due to the frequent clinical observation of reduced eye contact during social interactions. However, the paucity of neuroimaging techniques to acquire dyadic information on dynamic face processing during real social interactions has challenged advances in this field (Bolis & Schilbach, 2018; Rolison et al., 2015).

Conventional investigations of the neural underpinnings for face and social behaviors are based on single brain investigations primarily using neural imaging techniques such as functional magnetic resonance imaging (fMRI). Due to the single-subject conditions confined within one scanner bore, strict contraindications for head movement, severely isolated conditions, high magnetic field, and intense auditory distractions, the research focus has been on cognitive and perceptual processes within single brains without opportunities to investigate live and natural social interactions. These conventions have led to a long-standing experimental paucity of two-person interactive experimental paradigms and findings in social neuroscience. Increasing awareness of this knowledge gap has resulted in frequent calls for studies of two-person interactions (Hasson & Frith, 2016; Redcay & Schilbach, 2019; Schilbach et al., 2013).

As is frequently the case, advances in science are preceded by developments in technology. Recent developments in functional near-infrared spectroscopy (fNIRS) have been applied to hyperscanning (simultaneous brain scanning of two individuals during live interactions), to pave the way for much needed studies of live interactions between individuals and investigations of neural anomalies associated with interactive social deficits. Technical advances in fNIRS (Boas et al., 2014; Ferrari & Quaresima, 2012) and the immediate need to understand the biological components of live and interactive human social behaviors have supported the emergence of neuroimaging technology to investigate dynamic face-to-face and eye contact behaviors in ASD.

This investigation sought to to identify neural systems responsive to *in vivo* eye contact in ASD and examine their relationship to social symptomatology. The hypothesis was that interactive face processing, eye contact being a central component, engages complex social encoding due to high-level demands of rapid interpretation of subtle facial and eye movements that convey social meaning. Thus, it was expected that neural processing differences in ASD would include social systems (Carter & Huettel, 2013); interactive face processing systems (Hirsch et al., 2017; Kelley et al., 2021; Noah et al., 2020); and motion-sensitive systems (Braddick et al., 2003).

## Methods and Materials

Methods and materials are similar to those reported in a separate study currently under review (Hirsch et al., 2021). Hemodynamic signals were acquired for TD and ASD adults during live and interactive gaze at the eyes of a confederate and the eyes of a comparable dynamic video-face. In the real-person interaction condition (Real Eye), participants and a same-sex confederate viewed each other’s faces directly while sitting across a table. Findings were compared with the video condition in which participants and their confederate partners viewed the eyes of a size-matched face displayed on a video monitor (Video Eye). Eye-tracking during these conditions was confirmed compliance.

### Participants

Participants included 17 adults with ASD (3 female; mean age 25±4.9 years; 12 right-handed, 3 left-handed, and 2 ambidextrous (Oldfield, 1971)) whose diagnoses were verified by gold standard, research-reliable clinician assessments, including the Autism Diagnostic Observation Schedule, 2^nd^ Edition (ADOS-2 (Lord et al., 2012)) (Table 2), and expert clinical judgment using DSM-5 criteria (American Psychiatric Association, 2013); and 19 typically-developed (TD) adults (mean age 26±5.8 years; 18 right-handed and 1 ambidextrous) (Table 1). Participants were age and IQ matched (Tables 3 and 4) and recruited from ongoing research in the McPartland Lab, the Yale Developmental Disabilities Clinic, and the broader community through flyers and social media announcements. Inclusion criteria included age 18-45 years, IQ≥70, and English speaking. Exclusion criteria included diagnosis of bipolar disorder, personality disorder, or schizophrenia spectrum disorder; anti-epileptic, barbiturate, or benzodiazepine medication use; history of seizures, brain damage, or recent serious concussion; alcohol use within 24 hours; recreational drug use within 48 hours; chronic drug abuse; medication changes within two weeks; sensory impairment or tic disorder that would interfere with fNIRS recording; history of electroconvulsive therapy; or genetic or medical condition etiologically related to ASD. Additional exclusionary criteria for TD participants included self-report of any psychiatric diagnosis or learning/intellectual disability; psychotropic medication; or a first-degree relative with ASD. All participants provided written and verbal informed consent in accordance with guidelines and regulations approved by the Yale University Human Investigation Committee (HIC #1512016895) and were reimbursed for their participation. Assessment of the ASD participants’ capacity to capacity to give informed consent was provided by clinical research staff who monitored the process and confirmed verbal and non-verbal responses. ASD participants were accompanied at all times by a member of the clinical team, who continuously evaluated their sustained consent to participate.

**Table 1.** TD Study Participants and Behavioral Test Scores Demographic information for Typically-Developed (TD) participants. Assessment measures include the Wechsler Abbreviated Scale of Intelligence (WASI); Autism-Spectrum Quotient test (AQ); Broad Autism Phenotype Questionnaire (BAPQ); Social Responsiveness Scale (SRS); Beck Anxiety Inventory (BAI); State-Trait Anxiety Inventory (STAI); and the Liebowitz Social Anxiety Scale (LSAS). The right column indicates whether or not eye-tracking data were acquired.

| ID | M/F | Age | WASI | AQ | BAPQ | SRS | BAI | STAI | LSAS | Eye-tracking |
| --- | --- | --- | --- | --- | --- | --- | --- | --- | --- | --- |
| 1 | Male | 18-22 | * | 12 | 77 | 52 | 10 | * | 23 | Yes |
| 2 | Male | 18-22 | 123 | 23 | 118 | 55 | 4 | 39 | 52 | Yes |
| 3 | Male | 28-32 | 113 | 6 | 85 | 9 | 4 | 26 | 24 | Yes |
| 4 | Male | 23-27 | * | * | 72 | 16 | * | * | * | Yes |
| 5 | Female | 23-27 | 129 | 6 | 70 | 20 | 1 | 25 | 23 | Yes |
| 6 | Female | 23-27 | 114 | 8 | 79 | 27 | 5 | 28 | 43 | Yes |
| 7 | Female | 28-32 | 103 | 1 | 53 | 6 | 3 | 21 | 22 | Yes |
| 8 | Male | 28-32 | * | 26 | 109 | 48 | 0 | 23 | 23 | Yes |
| 9 | Female | 18-22 | 79 | 32 | 121 | 64 | 5 | 37 | 44 | Yes |
| 10 | Female | 23-27 | 119 | 22 | 111 | 37 | 26 | 62 | 60 | Yes |
| 11 | Female | 28-32 | 107 | 14 | 88 | 36 | 2 | 29 | 63 | * |
| 12 | Male | 28-32 | 111 | 10 | 73 | 25 | 5 | 28 | 27 | * |
| 13 | Female | 23-27 | 126 | 14 | 93 | 43 | 10 | 38 | 12 | * |
| 14 | Male | 18-22 | * | 25 | 105 | 84 | 7 | 27 | 59 | Yes |
| 15 | Female | 23-27 | 122 | 7 | 79 | 12 | 0 | 26 | 32 | Yes |
| 16 | Male | 38-42 | * | 21 | 124 | 63 | 9 | 56 | 56 | Yes |
| 17 | Male | 18-22 | * | 19 | 96 | 45 | 0 | 22 | 45 | * |
| 18 | Male | 28-32 | 119 | 19 | 89 | 38 | 12 | 26 | 46 | Yes |
| 19 | Male | 38-42 | 121 | 10 | 73 | 31 | 0 | 20 | 18 | Yes |

\*Data unavailable

**Table 2.** ASD Study Participants and Behavioral Test Scores Demographic information for Autism Spectrum Disorder (ASD) participants. Assessment measures include the Wechsler Abbreviated Scale of Intelligence (WASI); Autism-Spectrum Quotient test (AQ); Broad Autism Phenotype Questionnaire (BAPQ); Social Responsiveness Scale (SRS); Beck Anxiety Inventory (BAI); State-Trait Anxiety Inventory (STAI); Liebowitz Social Anxiety Scale (LSAS); and the Autism Diagnostic Observation Schedule (ADOS). The right column indicates whether or not eye-tracking data were acquired.

| ID | M/F | Age | WASI | AQ | BAPQ | SRS | BAI | STAI | LSAS | ADOS | Eye-tracking |
| --- | --- | --- | --- | --- | --- | --- | --- | --- | --- | --- | --- |
| 1 | Male | 28-32 | 114 | 26 | 123 | 54 | 16 | 21 | 30 | 12 | Yes |
| 2 | Male | 33-37 | 104 | 23 | 121 | 30 | 8 | 31 | 0 | 15 | * |
| 3 | Male | 28-32 | 112 | 30 | 137 | 82 | 11 | 27 | 46 | 16 | * |
| 4 | Male | 23-27 | 108 | 15 | 102 | 53 | 21 | 30 | 56 | 14 | Yes |
| 5 | Male | 23-27 | 97 | 15 | 99 | 41 | 19 | 29 | 43 | 10 | Yes |
| 6 | Male | 23-27 | 90 | 26 | 131 | 132 | 41 | 53 | 79 | 16 | * |
| 7 | Male | 28-32 | 113 | 15 | 94 | 68 | 7 | 44 | 44 | 13 | * |
| 8 | Male | 18-22 | 95 | 24 | 117 | 79 | 24 | 37 | 38 | 15 | Yes |
| 9 | Male | 18-22 | 101 | 30 | 140 | 96 | 27 | 46 | 85 | 18 | Yes |
| 10 | Female | 28-32 | 110 | 20 | 124 | 70 | 9 | 61 | 34 | 17 | Yes |
| 11 | Female | 18-22 | 128 | 32 | 121 | 68 | 4 | 30 | 32 | 9 | Yes |
| 12 | Male | 18-22 | 124 | 25 | 128 | 60 | 1 | 35 | 44 | 9 | Yes |
| 13 | Male | 18-22 | 107 | 14 | 95 | 34 | 9 | 55 | 5 | 12 | Yes |
| 14 | Male | 18-22 | 121 | 30 | 155 | 103 | 17 | 41 | 77 | 11 | Yes |
| 15 | Male | 23-27 | 112 | 20 | 117 | 36 | 3 | 23 | 3 | 13 | Yes |
| 16 | Male | 28-32 | 101 | 47 | 189 | 153 | 25 | 67 | 126 | 12 | * |
| 17 | Female | 28-32 | 109 | 23 | 94 | 65 | 15 | 33 | 38 | 11 | Yes |

\*Data unavailable

**Table 3.**
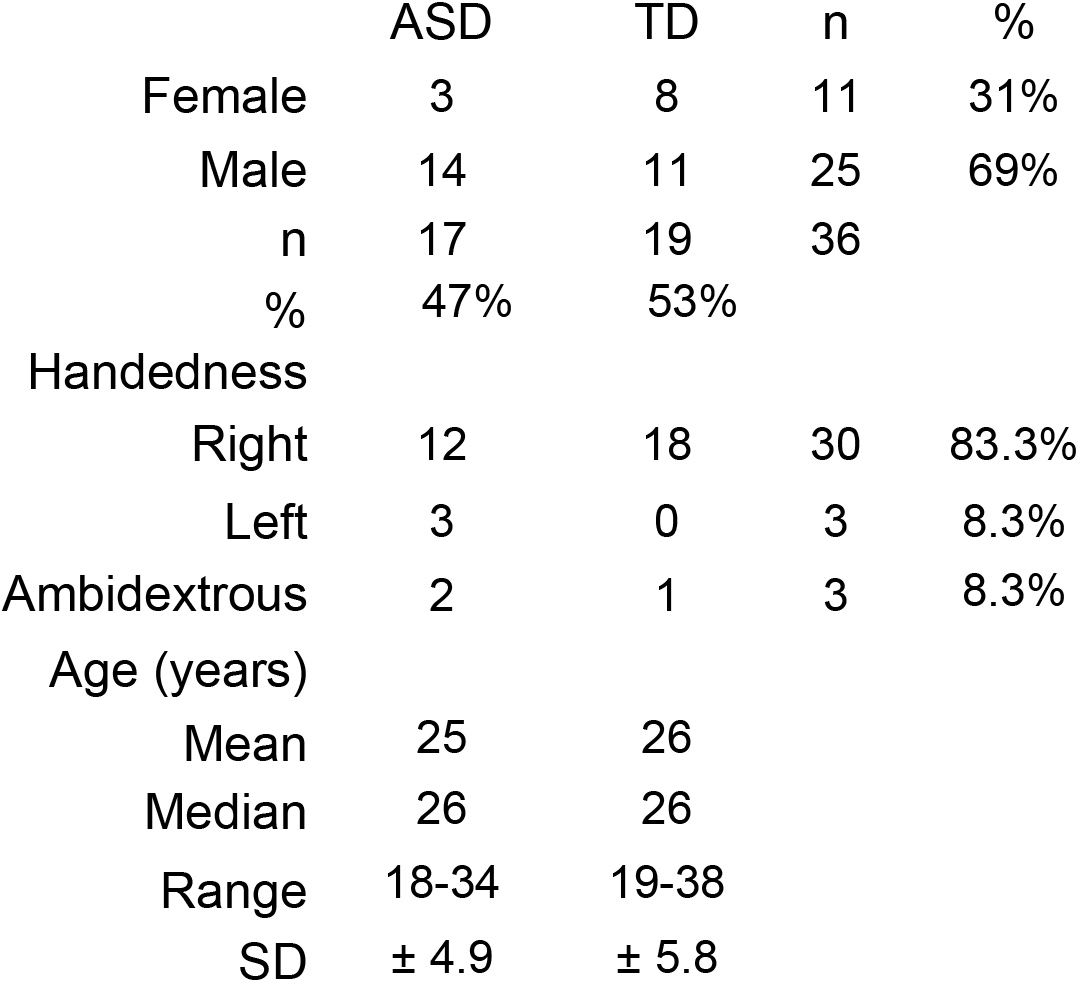
Comparison of ASD and TD Groups by Gender, Handedness, and Age Demographic details confirm similar distributions of gender, handedness, and age for the ASD and TD groups.

**Table 4.** Comparisons of Clinical and Behavioral Assessments for ASD and TD Groups Statistical comparisons (independent t-tests, two-tailed assuming unequal variances) of scores between Typically-Developed (TD) and autism spectrum disorder (ASD) groups are consistent with differences for the Autism-Spectrum Quotient test (p≤0.002), Broad Autism Phenotype Questionnaire (p≤0.002), Social Responsiveness Scale (p≤0.002), and Beck Anxiety Inventory (p≤0.016). No evidence was found for differences between the groups for the Wechsler Abbreviated Scale of Intelligence, State-Trait Anxiety Inventory, or the Liebowitz Social Anxiety Scale.

| Measure | p-value |
| --- | --- |
| Autism-Spectrum Quotient | 0.002 |
| Broad Autism Phenotype Questionnaire | 0.002 |
| Social Responsiveness Scale | 0.002 |
| Beck Anxiety Inventory | 0.016 |
| State-Trait Anxiety Inventory | 0.067 |
| Liebowitz Social Anxiety Scale | 0.363 |
| Wechsler Abbreviated Scale of Intelligence | 0.539 |

All participants were characterized by self-reported sex assigned at birth, age, full-scale IQ (FSIQ-4 as estimated by the Wechsler Abbreviated Scale of Intelligence, 2^nd^ Edition (WASI-II) (Wechsler, 2011), and self-reported clinical characteristics on several questionnaires, including the Autism-Spectrum Quotient (AQ (Baron-Cohen et al., 2001)); Broad Autism Phenotype Questionnaire (BAPQ (Hurley et al., 2007)); Social Responsiveness Scale, Second Edition (SRS-2 (Constantino & Gruber, 2012)); Beck Anxiety Inventory (BAI (Beck & Steer, 1991)); State-Trait Anxiety Inventory (STAI (Spielberger et al., 1983)); and the Liebowitz Social Anxiety Scale (LSAS (Fresco et al., 2001)). See Tables 3 and 4 for detailed demographic and statistical comparisons between the two groups. Group comparisons of clinical assessments indicated expected differences on the AQ (p≤0.01); BAPQ (p≤0.01); SRS (p≤0.01); and BAI scales (p≤0.05), and failed to provide evidence for differences on the WASI-II, STAI, and LSAS between the groups. Assessment and diagnostic tests were performed in clinical facilities at the Yale Child Study Center.

Participants were escorted from the clinical environment to the research environment for fNIRS / eye-tracking experiments. A clinical investigator was present during the data acquisition and monitored signs of discomfort during the experiment. All participants were paired with a same-gender TD confederate. One male and one female, both in their 20’s, served as confederates for all participants. Confederates were not informed of the participant’s diagnostic group membership before the experiment. Determination of a sample size sufficient for a conventional power of 0.80 is based on contrasts (Real face > Video Face) observed from a previous similar study (Noah et al., 2020). Using the power package of R statistical computing software (R Core Team, 2018) a significance level of 0.05 is achieved with 15 pairs. Sample sizes of 17 pairs (ASD) and 19 pairs (TD) assured adequate effect sizes.

#### Experimental Procedures and Paradigm

Dyads (participant and gender-matched confederate) were seated approximately 140 cm across a table from each other and set up with an extended head-coverage fNIRS cap and remote (table-top) eye-tracking. Each participant was instructed to look straight ahead either at their partner or at a monitor adjusted in size to subtend the same visual angles as the real face. In the live “Real Eye” task, dyads were instructed to gaze at each other’s eyes during cued 3-second epochs, and in the video (“Video Eye”) task, dyads were instructed to gaze at the eyes of the face as it appeared in a dynamic video). The video was a recorded version of a same-sex participant performing the same task while wearing the same optode cap as live participants.

For computational purposes, an “eye box” was constructed as a target region enclosing the eyes of the participants. This region subtended approximately 3.3 x1.5 deg of visual angle and defined the location of the region designated as the eye-to-eye contact zone for each participant. In both tasks, dyads alternated their gaze between the eyes of their (real or video) partner and two small light-emitting diodes (LEDs) located 10° to the left and 10° to the right of their partner.

The order of runs was randomly sequenced between viewing a real partner directly (Real Eye) or viewing a visual-angle corrected video partner (Video Eye) on a 24-inch 16×9 computer monitor placed back-to-back between participants, including a partition to assure that dyads could not see their real partner during video conditions. The face and distance of the video stimuli were calibrated to subtend identical degrees of visual angle in the field of view of the participants, and the timing and range of motion of eye movements between partners were the same in both tasks. The time-series (See Figure 1A) and experimental details included here for completeness were similar to prior studies (Hirsch et al., 2017; Noah et al., 2020).

**Figure 1.**
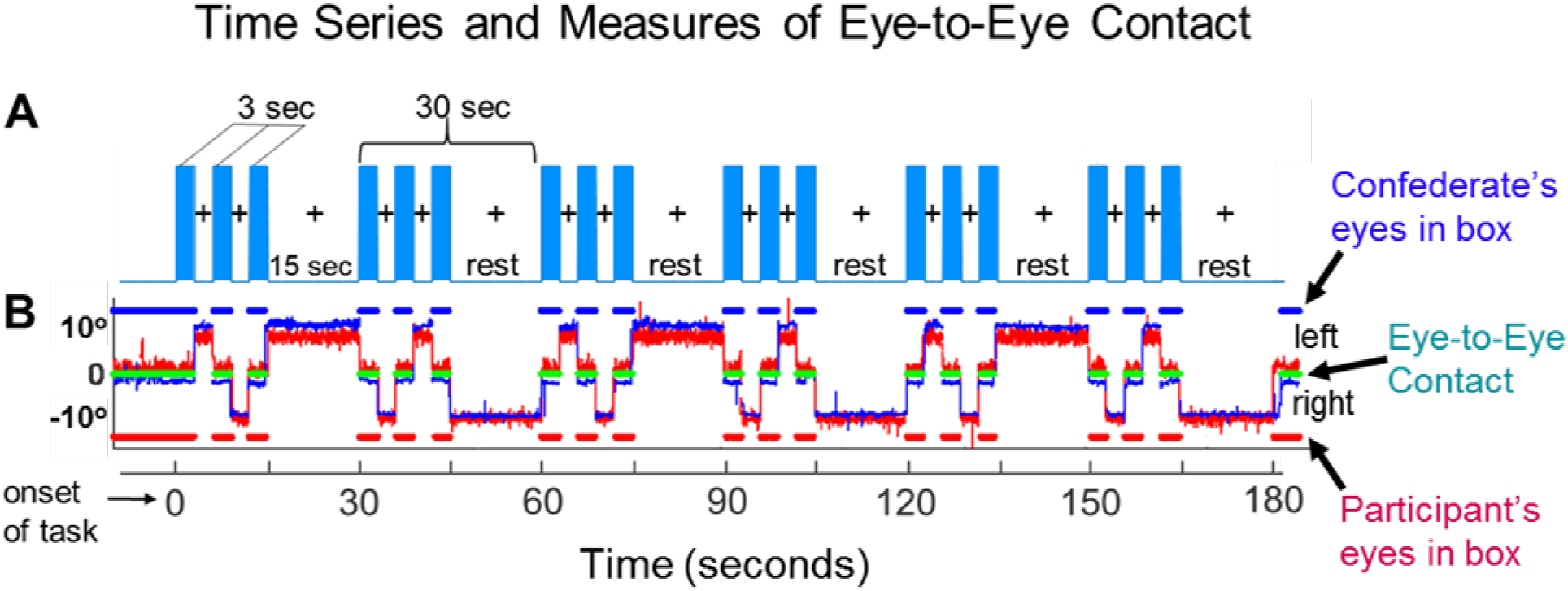
**A. Time course**. The duration of the run was three minutes, and each run was repeated twice for both the Real Eye and the Video Eye conditions. Each run included six alternating 15-second task and rest periods. In the task periods (blue bars), participants alternated their gaze in three-second epochs between the eyes of the confederate (Real Eye) or the person in the video (Video Eye) and the left or right lighted LED. During the 15-second rest period, participants looked only at the lighted LED. Gaze locations we confirmed by eye-tracking. **B. Eye-tracking: A single dyad illustration**. Red traces represent the eye movements of an ASD participant; blue traces represent the eye movements of a confederate partner. The eye-tracking data acquired on the Tobii system provides a frame-by-frame (8 ms) binary value that indicates whether or not eye gaze was directed within the eye-box of the partner. The blue dashed line (top) represents the duration of eye gaze (number of frames) that the confederate’s gaze was within the eye-box of the participant. Similarly, the red dashed line (bottom) represents the duration of gaze (number of frames) that the participant’s eye gaze was in the eye-box of the confederate. The green dashed line (middle) represents the length of time (number of frames) that the eyes of both partners were simultaneously focused within each other’s eye-boxes for a minimum of 83 ms. This is taken as a measure of eye-to-eye contact between the participant and the confederate.

At the start of each task, an auditory cue prompted participants to gaze at the eyes of their real or recorded partner. Subsequent auditory tones alternatingly cued eye gaze between eyes or LED according to the protocol time series. The 15-second active task period alternated with a 15-second rest/baseline period. The task period consisted of three 6-second cycles in which gaze alternated between the partner for 3 s and a lighted LED to either the right or left (alternating) of the participant for 3 s for each of three events. The time series was performed in the same way for all runs. The order of runs was counterbalanced across pairs of participants. During the 15-second rest/baseline period, participants focused on the lighted LED, as in the case of the 3-second periods that separated the eye contact and gaze events. The 15-second activity epoch with alternating eye contact events was processed as a single block.

The experimental paradigm employed a classic hemodynamic time series with 15 s of task alternating with 15 s of rest (Figure 1A). Run length was 3 m and included six task-rest cycles. Due to the social discomfort associated with prolonged mutual gaze at another’s eyes, the task epochs were subdivided into events (epochs) that alternated between three 3-second eye-on and 3-second eye-off cycles. During the “eye-on” epoch, dyads were instructed to gaze at the eyes of their (real or video) partner, making eye contact as often as possible in natural intervals. An auditory tone signaled the transition between eye-on and eye-off events indicating when participants were instructed to divert their gaze to the LED targets 10° to the right or left.

#### Eye Tracking

Two Tobii Pro x3-120 eye trackers (Tobii Pro, Stockholm, Sweden), one per participant, were used to acquire eye-tracking data at a sampling rate of 120 Hz. Eye trackers were mounted on the table surface facing each participant. Prior to the start of the experiment, a three-point calibration method was used to calibrate the eye tracker on each participant. The partner was instructed to stay still and look straight ahead while the participant was told to look first at the partner’s right eye, then left eye, then the tip of the chin. The same calibration procedure for video interactions was performed before recording on a still image presented on the monitor 70 cm in front of the participants. Similar “live-calibration” procedures have been used successfully in prior investigations of in-person social attention (Falck-Ytter, 2015; Thorup et al., 2016). As instructed for the eye-movement task, participants alternated their gaze between ≈0° and 10° of deflection. Participants fixated on the eyes of the video (Video Eye Condition) or the eyes of the confederate partner (Real Eye Condition) ±10° deflections to either the left or right. The eye contact portions of the task were 3 s in length, with six per trial, for 18 s of expected eye contact over the trial duration (Figure 1A).

Eye-tracking confirmed compliance with task instructions, as is illustrated in Figure 1B for a confederate (blue trace) and an ASD participant (red trace). The x-axis represents the run series (180 s), and the y-axis represents the gaze angle, where 0 represents eye-to-eye contact and ±10° indicates left and right deflections respectively. The moments of dyadic eye contact (gaze is within the eye-box of their partner) are indicated by the green line. The time series of Fig 1A and 1B are synchronized for illustrative purposes. The blue and red dashed lines above and below the eye position trace indicate the times of gaze locations that are within the eye-box of the partner for the confederate and the ASD participant respectively. An eye-to-eye contact event is defined as when the gazes of both partners are within the designated eye-box of the other for a minimum of 83 ms, 10 frames (Dravida et al., 2020). The green-colored dots in the figure indicate these 10-frame time points where the gaze of both partners was in the “eye-box” of the other. Eye contact performance estimates are shown in Figure 2 for an illustrative TD participant (Fig 2A) and an illustrative ASD participant (Fig 2B) on a trial by trial basis. The percentage of time in the eye-box of the confederate is represented by a color bar for the entire run time (180 s).

**Figure 2.**
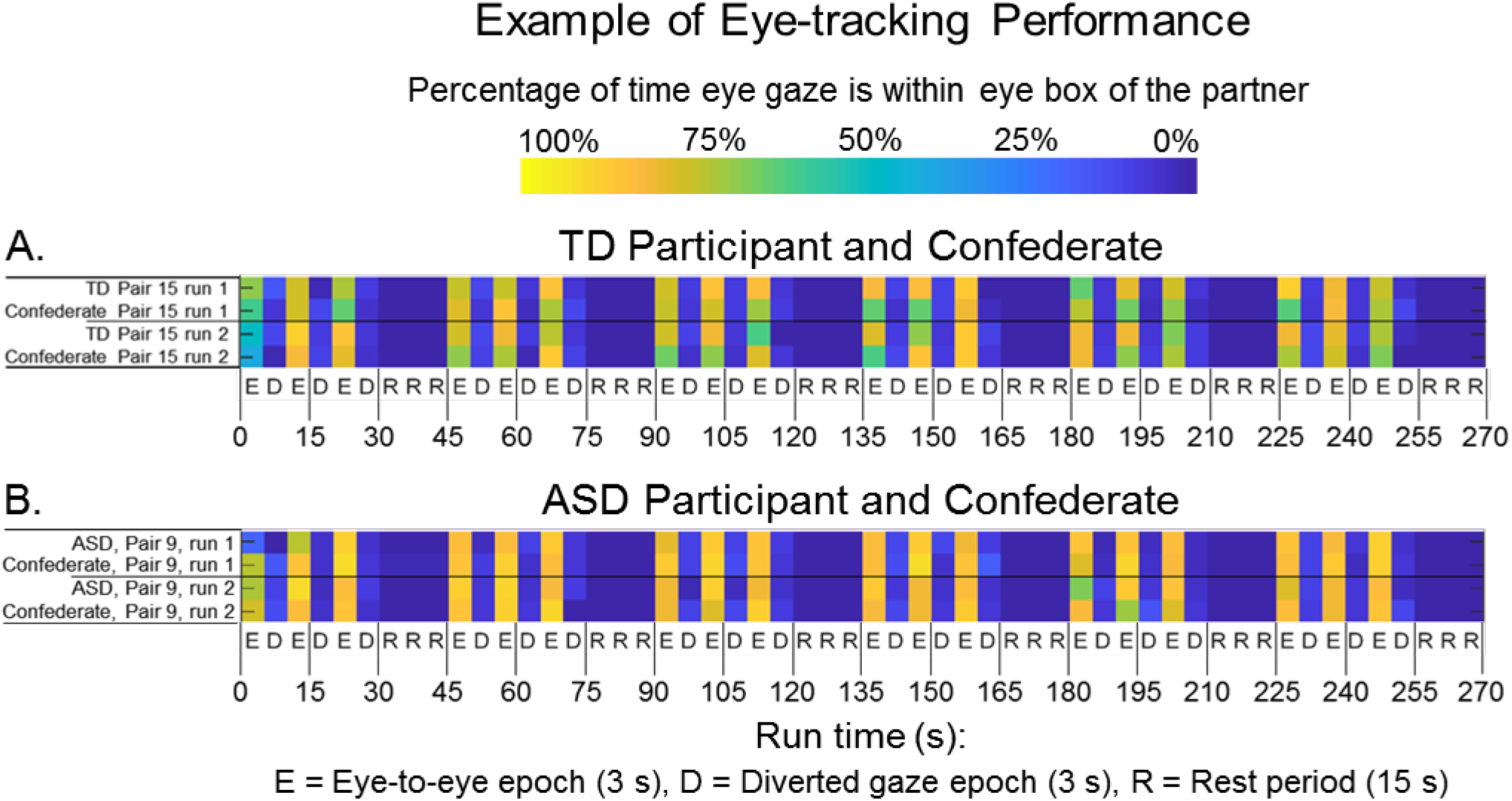
Gaze performance. Eye contact epochs (3 s) are indicated as E; Diverted gaze epochs (3 s) are indicated as D; and Rest periods (15 s) are indicated as R. The color bar (top) indicates the percent of time eye gaze was within the eye box of the partner. **A**. Example eye-tracking report for one Typically-Developed (TD) participant and confederate pair, Real Eye Condition. **B**. Example eye-tracking report for one ASD participant and confederate pair, Real Eye Condition. Similar computations were performed for all participants when eye-tracking data were acquired.

Heat maps of average eye-tracking behavior over an entire run are shown in Figure 3A for five illustrative participants.

**Figure 3.**
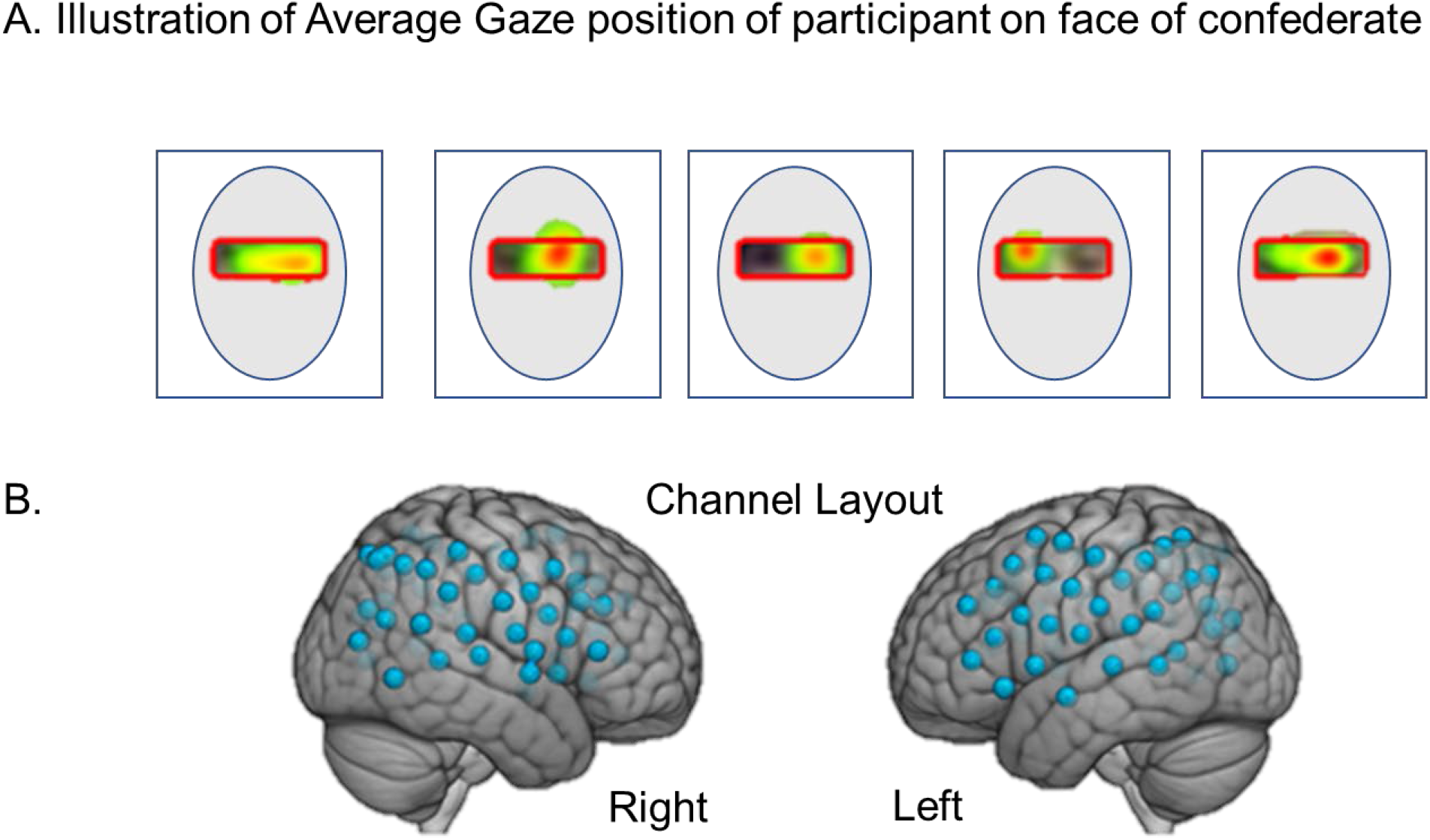
**A**. Examples of average participant eye-gaze positions when viewing the face of the confederate. The red box illustrates the target “eye box” and the color gradient from red to green indicates percent of “target hits” in the eye box for an entire run. **B**. Channel layout. Right and left hemispheres of a single rendered brain illustrate median locations (blue dots) for 58 channels per participant. Montreal Neurological Institute (MNI) coordinates were determined for each channel by digitizing emitter and detector locations in relation to anterior, posterior, dorsal, and lateral fiduciary markers based on the standard 10-20 system.

#### Functional NIRS (fNIRS) Signal Acquisition and Channel Localization

Functional NIRS signal acquisition, optode localization, and signal processing, including global mean removal, were similar to methods described previously (Dravida et al., 2018; Hirsch et al., 2018; Noah et al., 2017; Noah et al., 2015; Piva et al., 2017; Zhang et al., 2017; Zhang et al., 2016) and are briefly summarized below. Hemodynamic signals were acquired using three wavelengths of light, and an 80-fiber, multichannel, continuous-wave fNIRS system (LABNIRS, Shimadzu Corp., Kyoto, Japan). Each participant was fitted with an optode cap with predefined channel distances. Three sizes of caps were used based on the circumference of the heads of participants (60 cm, 56.5 cm, or 54.5 cm). Optode distances of 3 cm were designed for the 60 cm cap and scaled equally to smaller caps. A lighted fiber-optic probe (Daiso, Hiroshima, Japan) was used to remove hair from the optode channel before optode placement.

Optodes consisting of 40 emitters and 40 detectors were arranged in a custom matrix, providing a total of 54 acquisition channels per participant. The specific layout of the optode channels is shown in Figure 3B. For consistency, the placement of the most anterior channel of the optode holder cap was centered 1 cm above nasion. To assure acceptable signal-to-noise ratios, resistance was measured for each channel prior to recording, and adjustments were made for each channel until all recording optodes were calibrated and able to sense known quantities of light from each laser wavelength (Noah et al., 2015; Ono et al., 2014; Tachibana et al., 2011). Anatomical locations of optodes relative to standard head landmarks were determined for each participant using a Patriot 3D Digitizer (Polhemus, Colchester, VT) (Eggebrecht et al., 2014; Eggebrecht et al., 2012; Ferradal et al., 2014; Okamoto & Dan, 2005; Singh et al., 2005). Montreal Neurological Institute (MNI) coordinates (Mazziotta et al., 2001) for each channel were obtained using NIRS-SPM software (Ye et al., 2009).

#### Analysis of eye-tracking data

Eye-tracking data were exported from the Tobii system to the data processing pipeline and custom scripts in MATLAB were used to calculate the mutual eye contact events. Data were not usable on 5 out of 17 ASD participants and 4 out of 19 TD participants due to either calibration or equipment problems (right columns of Tables 1 and 2 summarize the eye-tracking acquisitions). Tobii Pro Lab software (Tobii Pro, Stockholm, Sweden) was used to create the eye-box as a target area of interest for subsequent eye-tracking analyses run in MATLAB 2014a (Mathworks, Natick, MA). The eye-tracking data for each run and each participant for both live and video sequences confirmed task compliance using both the trial by trial and the run average approaches.

#### fNIRS Signal Processing

Raw optical density variations were acquired at three wavelengths of light (780 nm, 805 nm, 830 nm), which were translated into relative chromophore concentrations using a Beer-Lambert equation (Hazeki & Tamura, 1988; Hoshi, 2003; Matcher et al., 1995). Signals were recorded at 30 Hz. Baseline drift was removed using wavelet detrending provided in NIRS-SPM (Ye et al., 2009). In accordance with recommendations for best practices using fNIRS data (Yücel et al., 2021), global components attributable to blood pressure and other systemic effects (Tachtsidis & Scholkmann, 2016) were removed using a principal component analysis (PCA) spatial global mean filter (Zhang et al., 2017; Zhang et al., 2016) before general linear model (GLM) analysis. All analyses are reported using the combined OxyHb and deOxyHb signals. The deOxyHb signal was inverted so that a positive result corresponded to increases in brain activity, similar to the OxyHb signal. The combined signal averages were taken as the input to the second level (group) analysis. Comparisons between conditions were based on GLM procedures using the NIRS-SPM software package. Event epochs within the time series were convolved with the hemodynamic response function provided from SPM8 (Penny et al., 2011) and were fit to the signals, providing individual “beta values” for each participant across conditions. The median “beta-value” represents the extent to which the signals in a cluster fit the model of the hemodynamic signal convolved with the time series. Group results based on these median beta values were rendered on a standard MNI brain template (ICBM152 T1 MRI template (Mazziota et al., 2001) in SPM8 using NIRS-SPM software (Ye et al., 2009). Anatomical correlates were estimated with the TD-ICBM152 atlas using WFU PickAtlas (Maldjian et al., 2004; Maldjian et al., 2003).

#### Code Accessibility

**C**ustom code will be provided upon request at fmri.org.

## Results

### ADOS scores and neural responses: Neuroimaging

Neural responses (beta values) acquired during eye-gaze were regressed by the individual ADOS scores using general linear models. The whole-brain main-effect of the eye contact activity related to ADOS scores is shown in Figure 4. Blue clusters indicate regions of the brain where neural activity as represented by the individual average was negatively related to the individual ADOS scores. That is, participants with higher ADOS scores showed consistently lower live eye contact related neural activity located in the right dorsal stream, including angular gyrus (AG), supramarginal gyrus (SMG), somatosensory association cortex (SSAC), and somatosensory cortex (SSC), and also the right dorsolateral prefrontal cortex (DLPFC) (p < 0.01). See Table 5.

**Figure 4.**
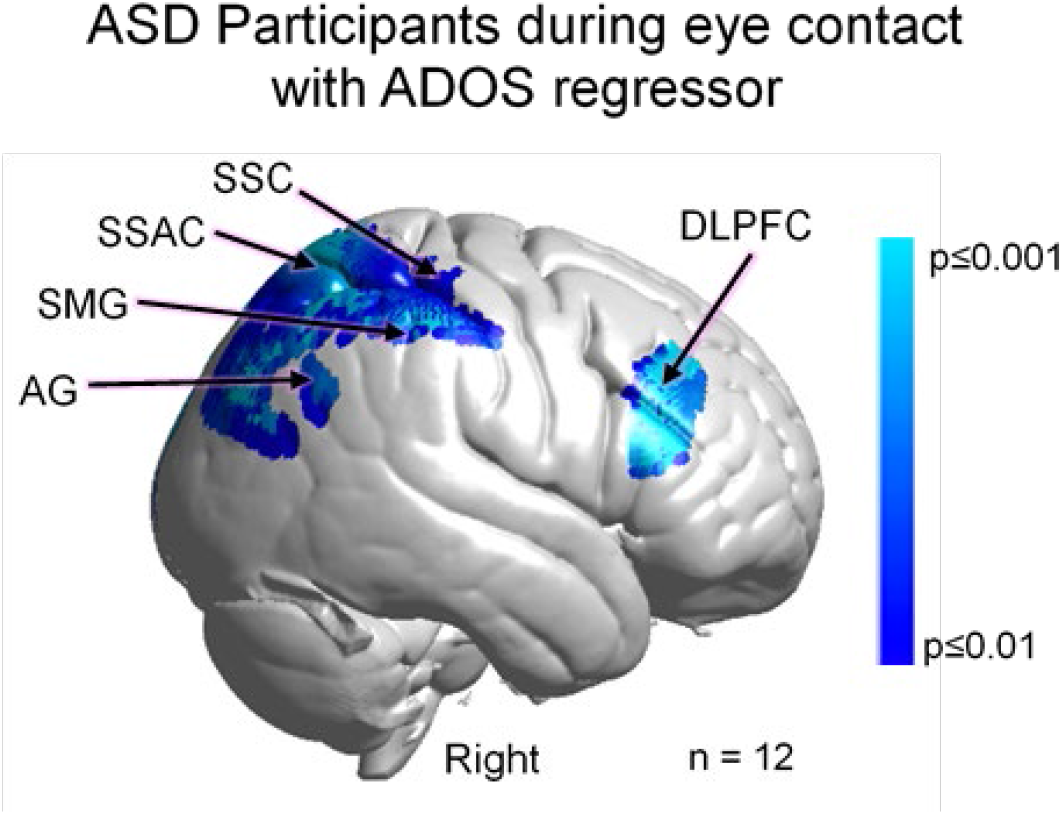
Main effect neural results for Autism Spectrum Disorder (ASD) participants during real eye contact with ADOS (Autism Diagnostic Observation Schedule) scores as a linear regressor. OxyHb and deOxyHb signals are combined. (See Table 5). Blue colors indicate a negative relationship between neural responses and ADOS scores indicating that as symptom severity increases neural responsiveness in these regions decreases. Light blue indicates responses corrected for multiple comparisons using FDR at p≤0.01. SSC: somatosensory cortex; SSAC: somatosensory association cortex; SMG: supramarginal gyrus; and AG: angular gyrus.

**Table 5.**
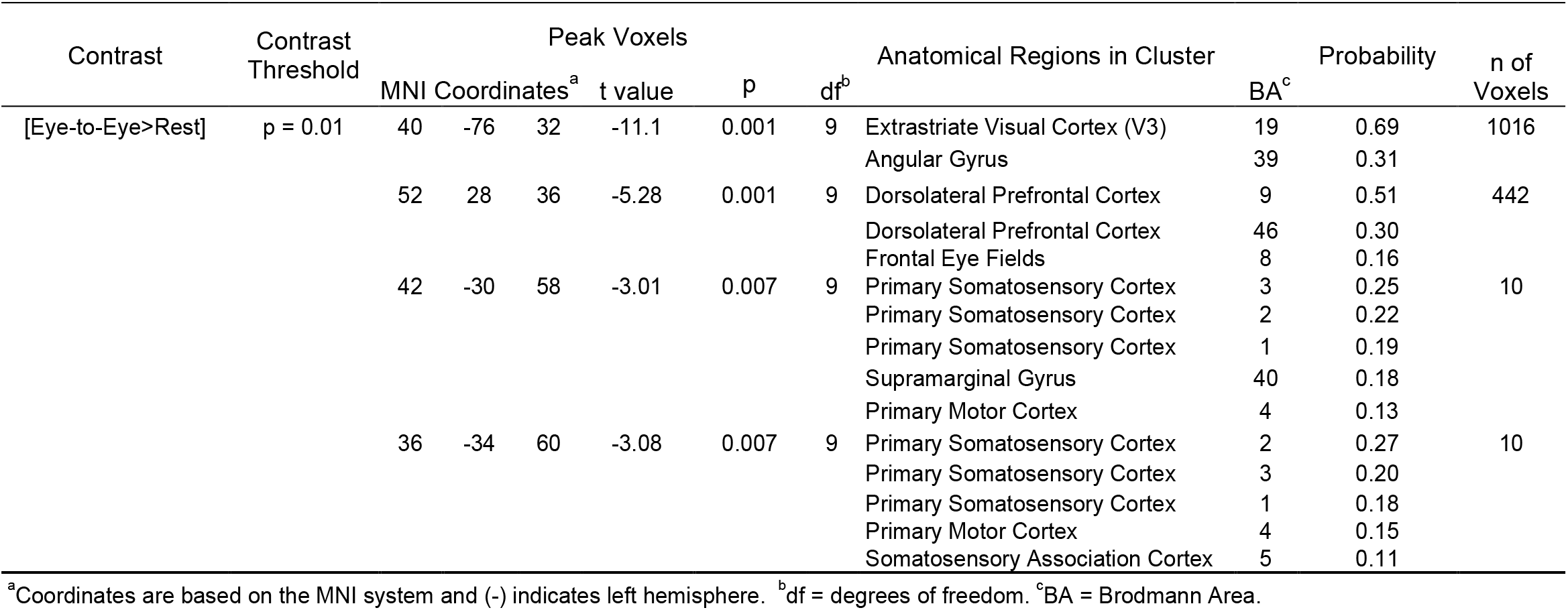
GLM Contrast comparison: [Eye-to-Eye>Rest] with ADOS regressor (deOxyHb + OxyHb signals)

### ADOS scores and neural responses: Individual Differences

The individual ADOS scores for each participant (identified by participant number in Table 2) are plotted against the median “beta-values” of the fNIRS signal (Figure 5). The best fit line illustrates the negative relationship (r= -0.69) for the posterior regions and r=-0.77 for the anterior frontal area.

**Figure 5.**
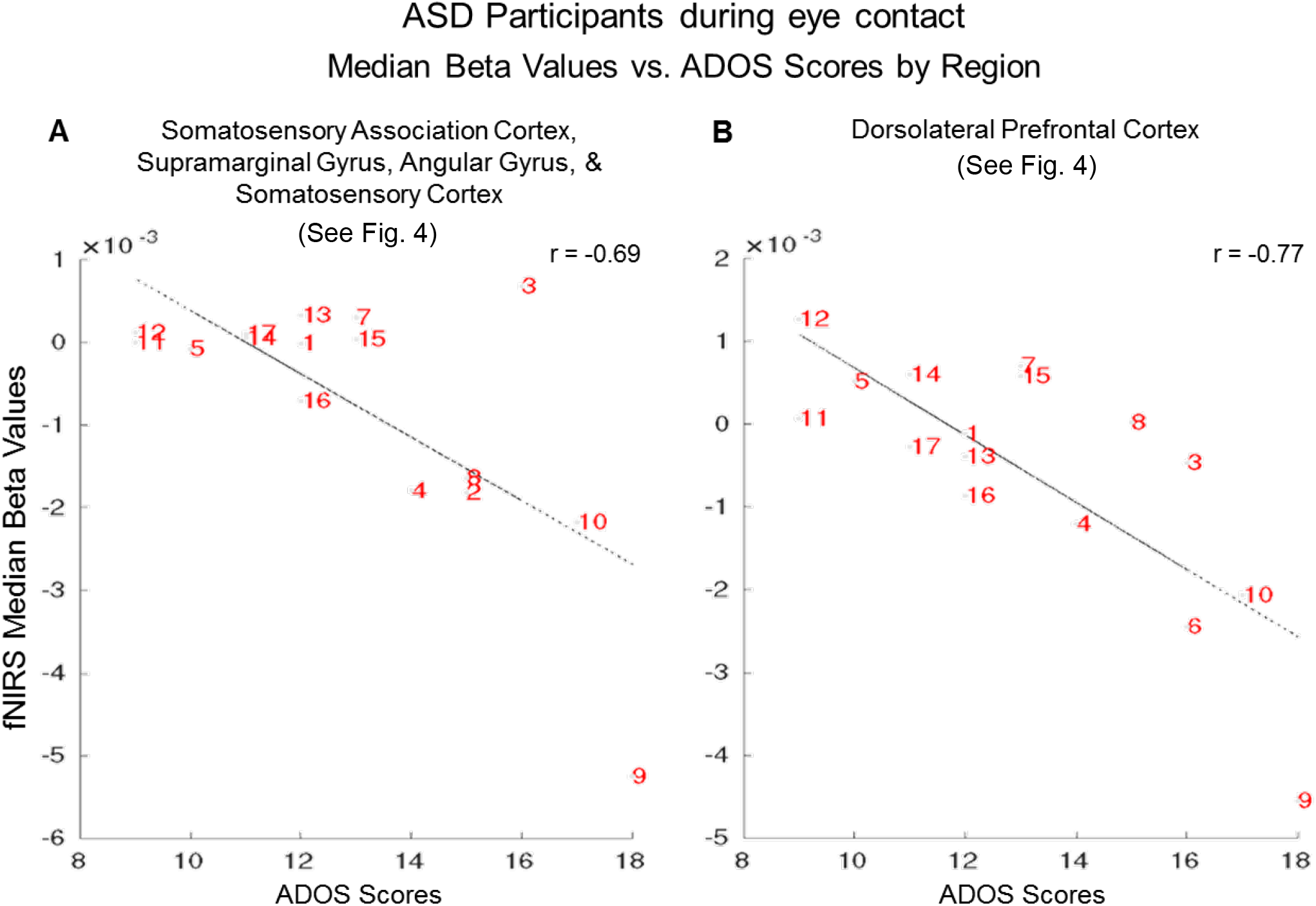
Autism Spectrum Disorder (ASD) participants (numbers correspond to Table 2) during eye contact “beta values” vs. ADOS (Autism Diagnostic Observation Schedule) scores. The median hemodynamic signals (Beta values, y-axis) within the responsive brain regions (Figure 4 and Table 5) and ADOS scores (x-axis) are shown for each participant. The main effect of eye-to-eye contact is negatively correlated with fNIRS signals in **A**. right hemisphere somatosensory association cortex, supramarginal gyrus, angular gyrus, and (r = -0.69); and somatosensory cortex **B**. right dorsolateral prefrontal cortex (r = -0.77).

### SRS scores and neural responses: Neuroimaging

To further evaluate the relationship between social symptomatology and live-face eye-gaze we combine the SRS scores for both ASD and TD groups based on the assumption that ASD traits are also present in the general population. The above findings presented in Figures 4 and 5 and Table 5 predict a similar relationship. Consistent with this prediction, a negative relationship (r = -0.58) was observed in regions located in the right dorsal stream including somatosensory cortex (SSC), somatosensory association cortex (SSAC), and supramarginal gyrus (SMG). The whole-brain analyses of the eye contact main effect was regressed by the SRS scores and shown in Figure 6 (blue clusters).

**Figure 6.**
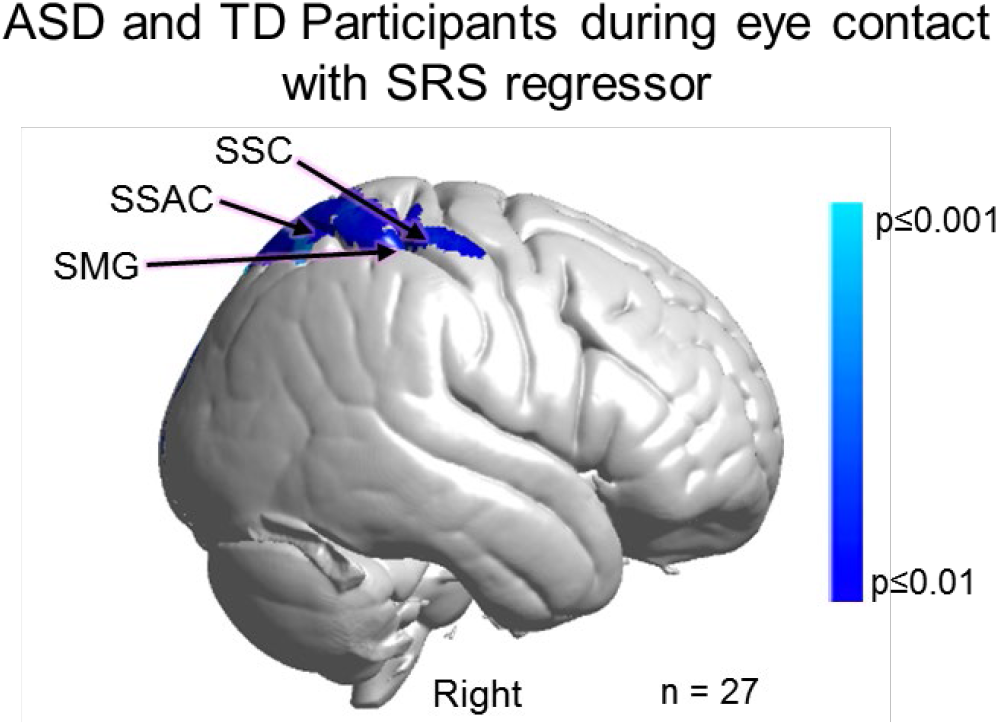
Main effect neural results for Autism Spectrum Disorder (ASD) and typically-developed (TD) participants during eye contact with SRS (Social Responsiveness Scale) scores as a linear regressor. OxyHb and deOxyHb signals are combined. Blue colors indicate a negative relationship between neural responses and SRS scores, which suggests that increased symptom severity is associated with reduced regional neural responsiveness. See Table 6. Light blue indicates responses corrected for multiple comparisons using FDR at p≤0.01. SSC: somatosensory cortex, SSAC: somatosensory association cortex, and SMG: supramarginal gyrus.

Participants with higher SRS scores indicating higher levels of autistic traits showed reduced neural activity during eye contact in the right somatosensory cortex (SSC), somatosensory association cortex (SSAC), and supramarginal gyrus (SMG) (p≤0.01). Table 6 provides the peak MNI coordinates, cluster t-values, anatomical regions within the cluster, Brodmann’s Area (BA), probability of occurrence in the cluster, and relative size of the active area (n of voxels).

**Table 6.**
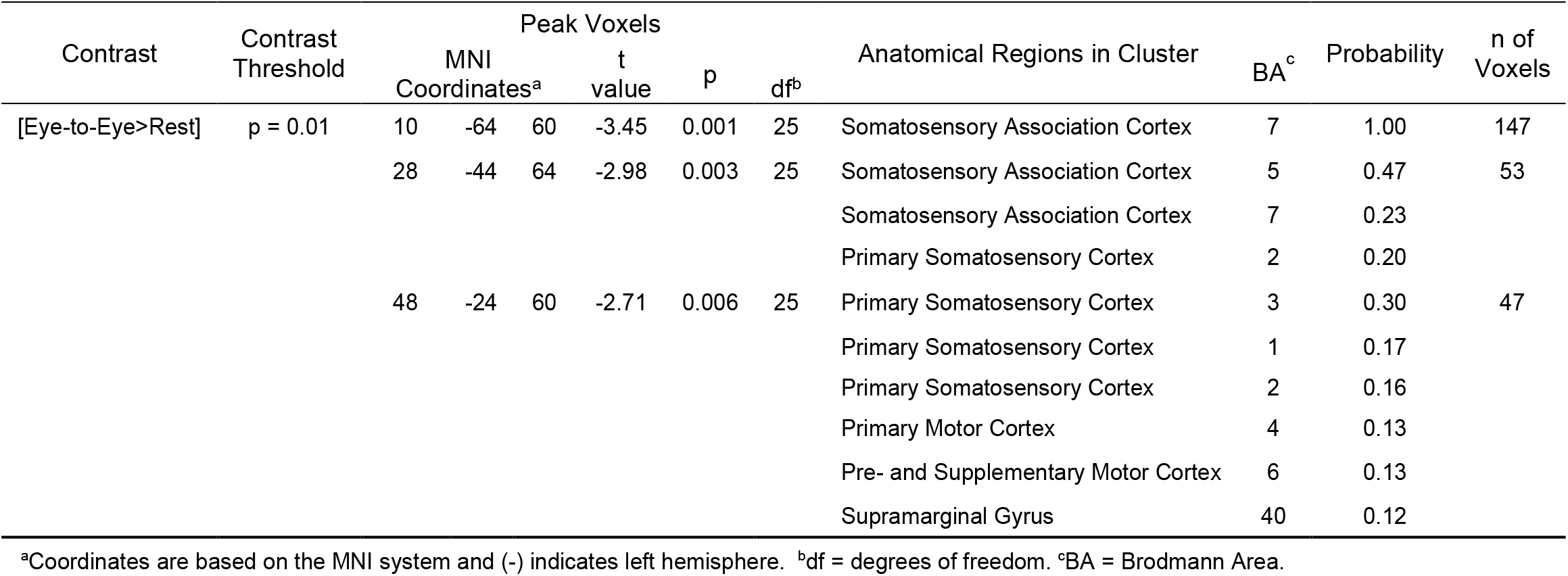
GLM Contrast comparison: [Eye-to-Eye>Rest] with SRS regressor (deOxyHb + OxyHb signals)

### SRS scores and neural responses: Individual Differences

The individual SRS scores for each participant (identified by a number that corresponds to the participant number in Tables 1 and 2) are plotted against the individual median beta-values of the fNIRS signal (Figure 7).

**Figure 7.**
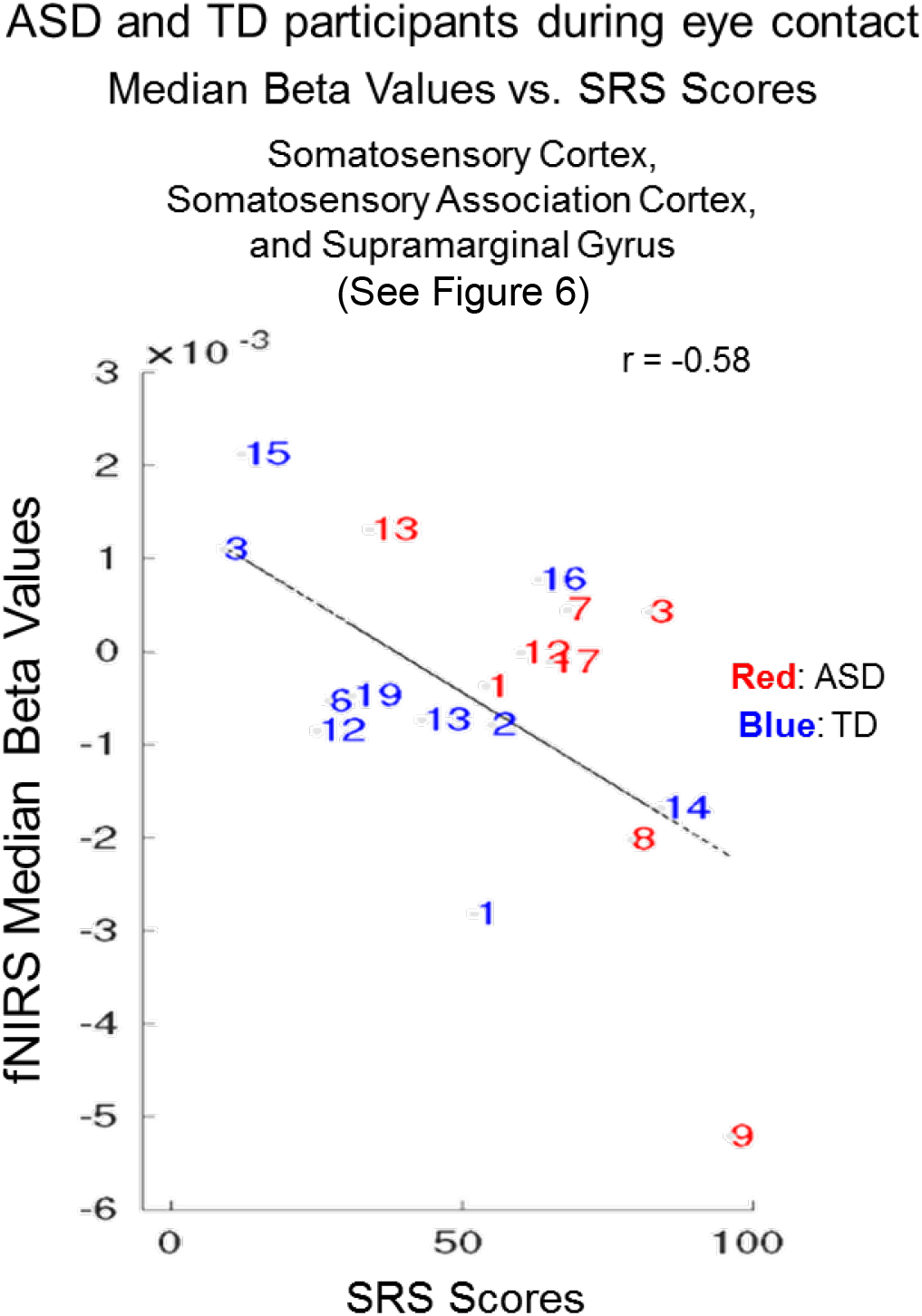
Autism Spectrum Disorder (ASD) participants (red numbers) and typically-developed (TD) participants (blue numbers) during eye contact vs. Social Responsiveness Scale (SRS) scores. The median hemodynamic signals (Beta values, y-axis) within the responsive brain region (Figure 6 and Table 6) and SRS scores (x-axis) are shown for each participant. The main effect of eye-to-eye contact is negatively correlated with fNIRS signals in right hemisphere somatosensory cortex, somatosensory association cortex, and supramarginal gyrus (r = -0.58). Numbers indicate individual participants shown in Table 2 (ASD participants, red numbers) and Table 1 (TD participants, blue numbers).

Blue numbers represent TD participants and red numbers represent ASD participants. The interspersal of the individual scores between ASD (red) and TD (blue) participants is consistent with the assumption that social responsiveness traits vary within the general population as well as within ASD. The best fit line illustrates the negative relationship (r= -0.58).

## Discussion

This application of two-person neuroimaging technology to investigate the relationship between the neural underpinnings of interactive face and eye contact and the severity of symptomatology in ASD addresses a prominent and understudied question of social deficits in ASD. Individual clinical evaluations of ASD symptom severity applied as a second level regressor on whole-brain neuroimaging findings acquired during live real person eye-to-eye contacts confirm a negative relationship between scores and neural signals in brain regions responsive to real eye-to-eye contacts. Participants with higher ADOS scores showed lower neural signals (beta-values, an indicator of signal strength and fit to the general linear model) in brain regions previously associated with social activity, interactive face processing, and motion sensitivity. Findings also included the right dorsolateral prefrontal cortex, a region implicated in both ASD and commonly co-occurring conditions, such as major depressive disorder. Further, a similar finding was observed for the SRS when the scores of both TD and ASD participants were combined for regions within the dorsal stream but the DLPFC. That is, as individual symptom severity increased as indicated by the elevated SRS, the neural signal decreased in the right dorsal-stream regions. A similar finding was observed for SRS scores and the relationship to neural signals acquired by fMRI in the fusiform gyrus and the amygdala during static face processing (Scherf et al., 2015). Interestingly, in this study, the SRS finding included TD as well as ASD participants suggesting that variations in social responsiveness and the associated reduction in dorsal stream activity are similarly represented in the general population.

Unique features of this study include the live interactive face task as well as the eye-tracking documentation of compliance combined with the individual difference approach to characterize single participants by both measures. All data were acquired during the epochs when participants directed their eye gaze to within the confederate eye box. Continuous monitoring confirmed high levels of compliance in all cases. That is, when asked to do the task, participants were able to do it although eye-to-eye contact was not necessarily a comfortable task for them.

These findings of a negative association between right dorsal regions and symptom severity do not imply a causal role between neural substrates and symptoms. However, it can be concluded that the dorsal regions found to be related to symptomatology (right hemisphere angular gyrus, supramarginal gyrus, somatosensory association cortex, somatosensory cortex, and dorsolateral prefrontal cortex), are vulnerable to the underlying neural conditions relevant to ASD. Given a well-recognized need for biomarkers for ASD that associate with the clinical phenotype at the individual level, the strong relationships observed between neural activity and both clinician and self-reported social function suggest potential utility in key contexts of use, such as stratification for enrichment of clinical trials (McPartland, 2017).

In conclusion, these findings highlight the right dorsal stream system and interactive face processing as a regions and tasks of interest for understanding the underlying neural deficits that distinguish ASD and TD participants. The specificity of these findings opens new directions for investigating these brain-to-behavior linkages. For example, these regions have previously been implicated in motion sensitivity (Braddick et al., 2003) and raise the interesting hypothesis that reduced face processing in social interactions in ASD is related to reduced sensitivity to the subtle expressive movements of a real face. However, the strong (r = -0.77) negative correlation between symptom severity (ADOS) and the dorsolateral prefrontal cortex in ASD does not fall under that hypothesis and was not predicted. These findings, however, are consistent with face processing observed right lateral prefrontal cortex using fMRI and TD participants (Chan & Downing, 2011). Interestingly, it was found in that study that face processing in the right inferior frontal junction (which includes regions labeled as DLPFC and frontal eye-fields in this study, see Table 5) was primarily responsive to the eyes and not the whole face. In this study the strongest negative correlation between ADOS scores and neural activity (−0.77) was in this area and observed during eye contact (Figure 4). These findings suggest another target of further investigation in this dorsal neural pathway and its role in the social symptomatology.

## Data Availability

The datasets analyzed for this study will be made available upon request at fmri.org/ ENDAR and the NIH Data Archive.

http://www.fmri.org

## Acknowledgements

This research was partially supported by the National Institute of Mental Health of the National Institutes of Health under award numbers 1R01MH111629 (PIs JH and JCM); R01MH107513 (PI JH); 1R01MH119430 (PI JH); U19MH108206 (PI JCM); R01 MH107426 (PI JCM); R01 MH100173 (PI JCM). The content is solely the responsibility of the authors and does not necessarily represent the official views of the National Institutes of Health. All data reported in this paper are available upon request from the corresponding and first author. The authors are grateful to the participants for their essential efforts to advance understanding of ASD; to our two confederates, CD and IS, for consistent partnership with our participants and the investigators; and to Jen Cuzzocreo for data-base management and graphical representations of the data.

## Disclosures

The authors declare that this research was conducted in the absence of any commercial or financial relationships that could be construed as a potential conflict of interest.

## Data Availability Statement

The datasets analyzed for this study will be made available upon request at fmri.org/ENDAR and the NIH Data Archive.

## Notes

### Competing Interest Statement

The authors have declared no competing interest.

### Author Declarations

The Human Research Protection Program of Yale University gave ethical approval for this work.

